# Using the shock index to rule out stroke in patients with impaired consciousness in the emergency department: a single-center retrospective study

**DOI:** 10.1101/2020.05.16.20101204

**Authors:** Kosuke Kato, Shogo Sagisaka, Masayuki Ueda, Hidetaka Tamune

**Author notes:** Corresponding authors: Kosuke Kato, Hidetaka Tamune.

## Abstract

**Background:** Stroke is a significant cause of impaired consciousness. During a stroke, intracranial pressure increases, resulting in hypertension and bradycardia, also known as the Cushing response. Previous studies showed that systolic blood pressure (sBP) was useful for diagnosing stroke. We hypothesized that stroke patients would show a low Shock Index (SI, pulse rate divided by sBP), and that a high SI would therefore be also useful for ruling out stroke in patients with impaired consciousness.

**Methods:** This was a single-centre retrospective medical record review conducted between 2015 and 2016. We identified 194 patients with stroke and 588 patients without stroke. We included all out-patients with impaired consciousness who underwent cranial computed tomography (CT) and allocated them to the stroke (cerebral infarction, intracerebral haemorrhage, and subarachnoid haemorrhage) and non-stroke groups. For both the sBP and SI, we calculated the sensitivity and specificity of each cut-off value and compared their diagnostic value.

**Results:** The area under the receiver operating characteristic curve was 0.801 (95% Confidence Interval: 0.761–0.840) for the sBP and 0.764 (0.729–0.799) for the SI. For an sBP higher than 90 mmHg and an SI lower than 1, the sensitivity was 99.5% for both. The specificity was 10.9% and 15.1%, respectively, and the negative likelihood ratio was 0.045 and 0.034, respectively. Subgroup analyses suggested that predictive value of SI was enough high regardless of the subtype of stroke, patient’s past history of hypertension, or level of consciousness.

**Conclusion:** SI may be useful for excluding stroke in patients with impaired consciousness. This study suggested that when the pulse rate exceeds sBP, stroke can be ruled out. In these cases, we suggest that physicians focus on non-neurological examinations and treatments rather than diagnostic examinations for stroke.

## 1. Introduction

Stroke is one of the most significant causes of impaired consciousness. Immediate diagnosis and timely intervention are important in its management [1]. Thus, simple, accurate, timely, and cost-effective methods are necessary for appropriate triage in emergency settings [2]. The Cincinnati Prehospital Stroke Scale (CPSS) is a subset of neurophysical examinations, widely used to detect suspected stroke in prehospital settings [3]. However, detailed neurological examination in unconscious patients is often difficult due to their inability to cooperate with the physicians’ requests. In many institutions, patients with impaired consciousness routinely undergo cranial computed tomography (CT). However, various diseases, including infection, electrolyte imbalance, and metabolic disorders, also result in impaired consciousness. Hence, determining the likelihood of stroke is crucial to prioritize appropriate examinations and treatments.

Vital signs are easily accessible parameters in all settings and can be quickly evaluated in patients with impaired consciousness. Previous studies have shown that systolic blood pressure (sBP) is useful for predicting the presence of stroke in patients with impaired consciousness [4–6], and Ikeda et al. showed that a low sBP indicated a sufficiently low probability of stroke [4].

Increased intracranial pressure results in bradycardia with hypertension, which is known as the Cushing response [7]. In contrast, intravascular hypovolaemia results in tachycardia with hypotension. This is observed in other causes of impaired consciousness, including infection and metabolic disorders. Shock Index (SI, pulse rate divided by sBP) is known as a predictor of intravascular volume loss [8]. In principle, it is low in patients with stroke and relatively high when the cause of impaired consciousness is not stroke. Although previous studies have demonstrated the usefulness of the SI in evaluating the severity of haemorrhagic trauma [9], pneumonia [10], and stroke [11], no reports have shown its diagnostic value for patients with impaired consciousness.

The present study focuses on the diagnostic value of the SI in adult patients with impaired consciousness. We hypothesized that patients with stroke would show a low SI, and that a high SI might be useful for ruling out stroke in patients with impaired consciousness.

## 2. Methods

### 2-1. Study Setting

This study was conducted at the Tokyo Metropolitan Tama Medical Center, a 789-bed tertiary care teaching hospital in Tokyo, Japan. Approximately 35,000 out-patients, including 7,700 ambulance-borne patients, visit the emergency department (ED) of this institution annually. Cranial CT and MRI are readily available here.

### 2-2. Study design

This is a retrospective observational study based on the medical records in the ED. We initially screened the out-patient list of the ED for those who underwent plain cranial CT between November 1, 2015 and October 31, 2016. We then included patients with impaired consciousness based on a Glasgow Coma Scale (GCS) of less than 15, or when patients were understood to have had impaired consciousness based on the medical records [12].

Patients with trauma or suspected trauma were excluded, since this study focused on disorientation due to endogenous causes. We also excluded patients with cardiac arrest on arrival at the ED, and patients with no recorded vital signs.

In Tokyo, there is an emergency service protocol which involves identifying patients with a suspected stroke using the CPSS and transferring them directly to hospitals with a stroke care unit. Owing to this, CPSS-positive patients have been screened out before arriving at out emergency unit. Diagnosing CPSS-negative patients is assumed to be more difficult in terms of quickly differentiating stroke from other causes of impaired consciousness. In this study, we collected the data of CPSS-negative patients and focused on diagnosing stroke among them.

We collected data on the patients’ vital signs upon arrival at the ED, particularly their blood pressure and pulse rate. SI was then calculated using these data. A history of hypertension, diabetes mellitus, or hyperlipidaemia was identified on the basis of a review of the medical records or a history of the use of antihypertensive, hypoglycaemic, or lipid lowering agents. The previous history of stroke was determined by reviewing medical records. The final diagnosis was made on the basis of medical records and imaging results. We defined stroke as a cerebral infarction (CI), intracerebral haemorrhage (ICH), or subarachnoid haemorrhage (SAH). The majority of the patients with infarction were diagnosed with a head MRI, but MRI was omitted when the patient was in critical condition. We allocated eligible patients into either the stroke or non-stroke group and performed statistical analysis.

### 2-3. Statistical Analysis

The data were imported to EZR ver. 2.3 (R ver. 3.2.2 32bit) for statistical analysis [13]. Patients’ characteristics were evaluated using the t-test and one-way analysis of variance for numeric variables, and Fisher’s exact test for categorical variables. Receiver operating characteristic (ROC) curves were drawn for both the sBP and SI, to differentiate between the stroke and non-stroke groups. The area under the curve (AUC) was then calculated for each parameter. AUCs were tested by DeLong’s test.

We calculated the sensitivity and specificity for each cut-off value for both the sBP and SI and estimated 95% confidence intervals. We conducted analysis in the subgroup with a past history of hypertension. We also conducted subgroup analysis based on level of consciousness, using a GCS of 8 as the cut-off value.

### 2-4. Ethical Considerations

The study protocol was approved by the Research Ethics Committee, Tokyo Metropolitan Tama Medical Center (Approval number: 28–142). Informed consent was obtained from participants using an opt-out form on the website. This study complied with the Declaration of Helsinki and the STROBE statement.

## 3. Results

### 3-1. Participants

A total of 4,678 patients with cranial CT scans were identified. Out of these, 3,852 patients were excluded because of a history of trauma, cardiac arrest on arrival, clear consciousness, or lack of data on their level of consciousness. Another 44 patients were excluded because of lack of data regarding their vital signs. Among the remaining 782 patients, 194 patients (24.8%) were diagnosed with stroke. This group comprised 77 patients (9.8%) with CI, 82 patients (10.5%) with ICH, and 35 patients (4.5%) with SAH (**Fig. 1**). The diagnoses of the non-stroke group patients are shown in **Table 1**.

**Fig 1.**
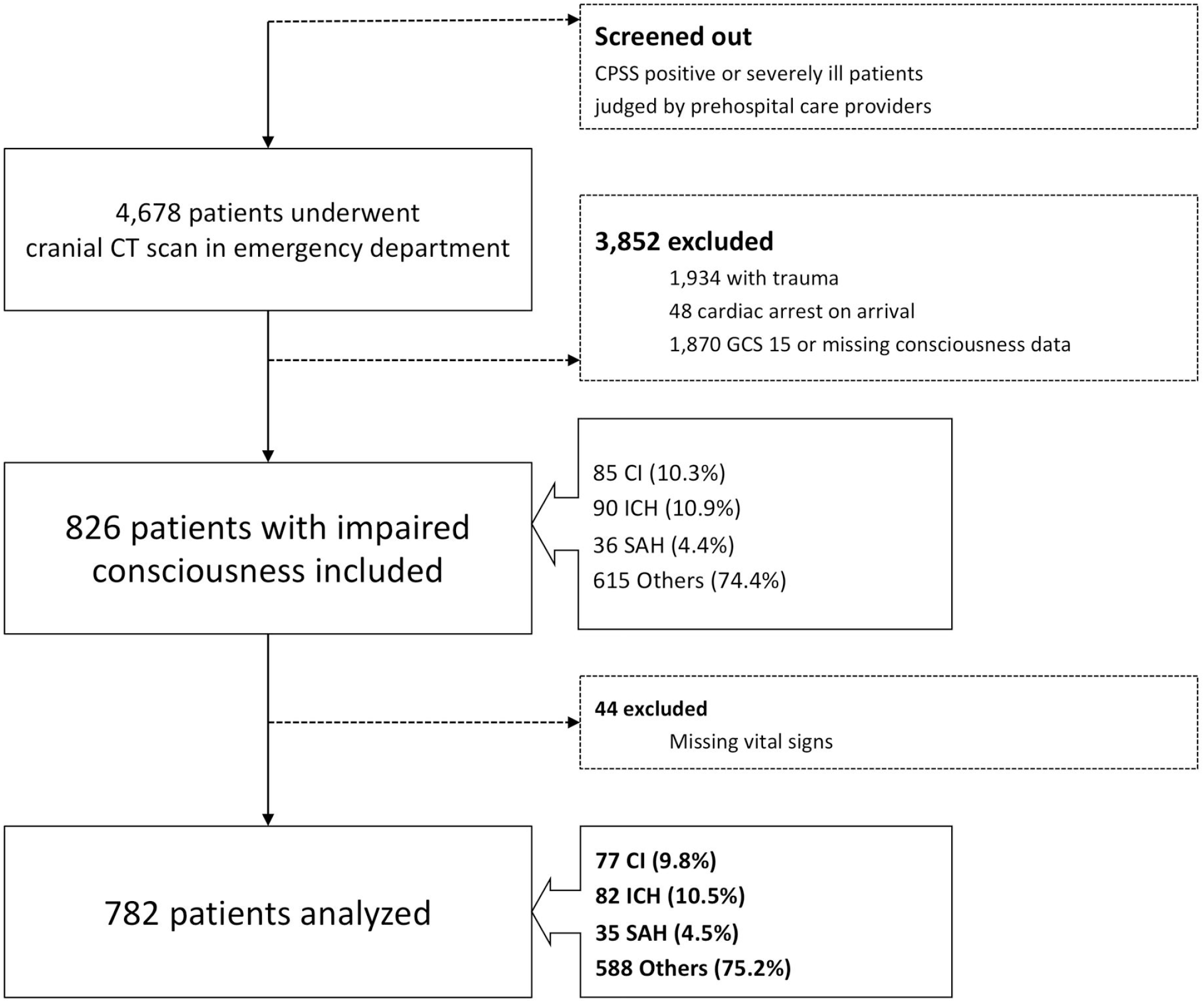
Flow chart of the study. A total of 4,678 CPSS-negative patients with cranial CT scan were identified. Exclusion criteria and distribution of the final diagnoses are shown. For more details, see the main text and Tables.

**Table 1.**
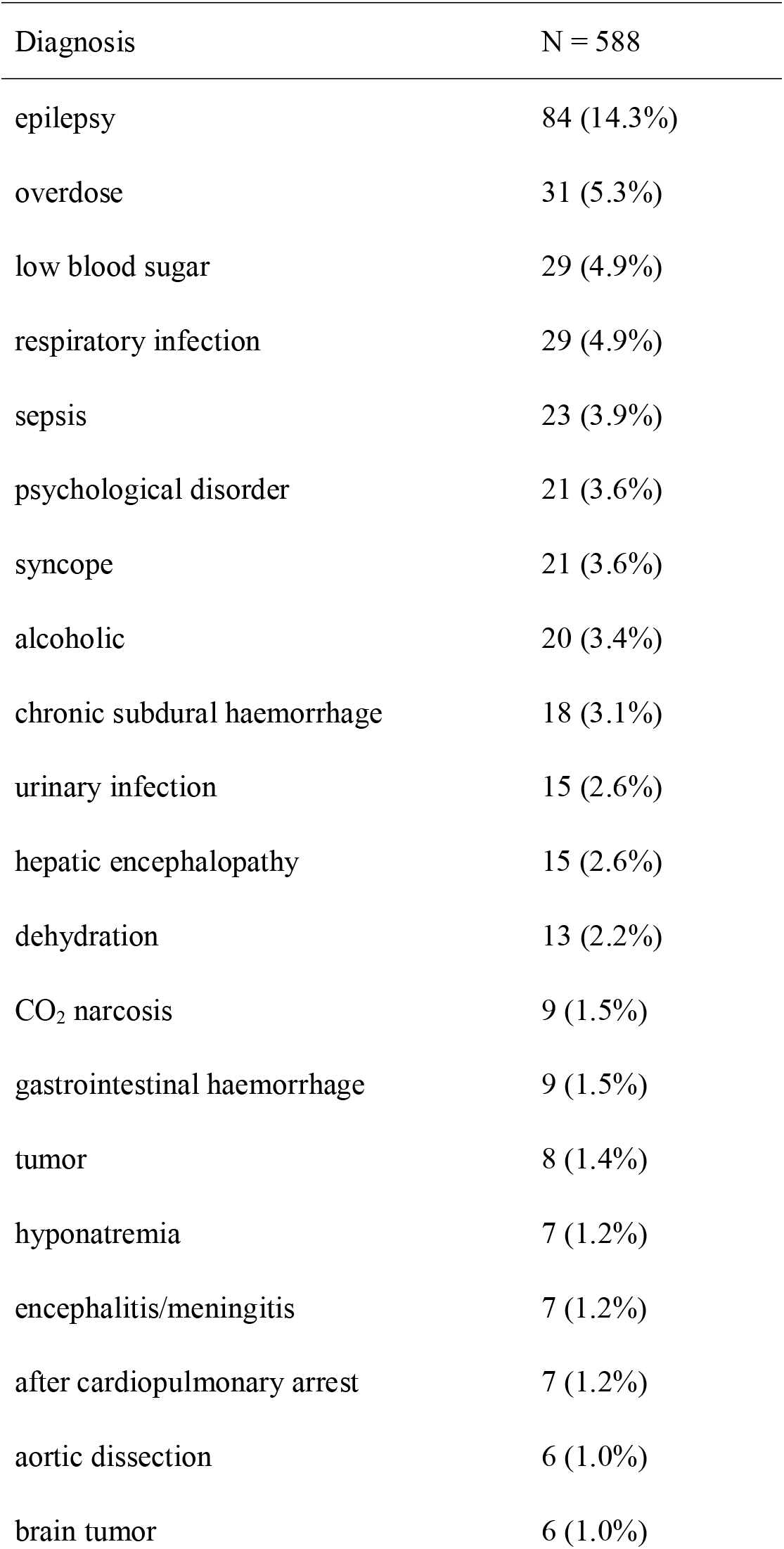
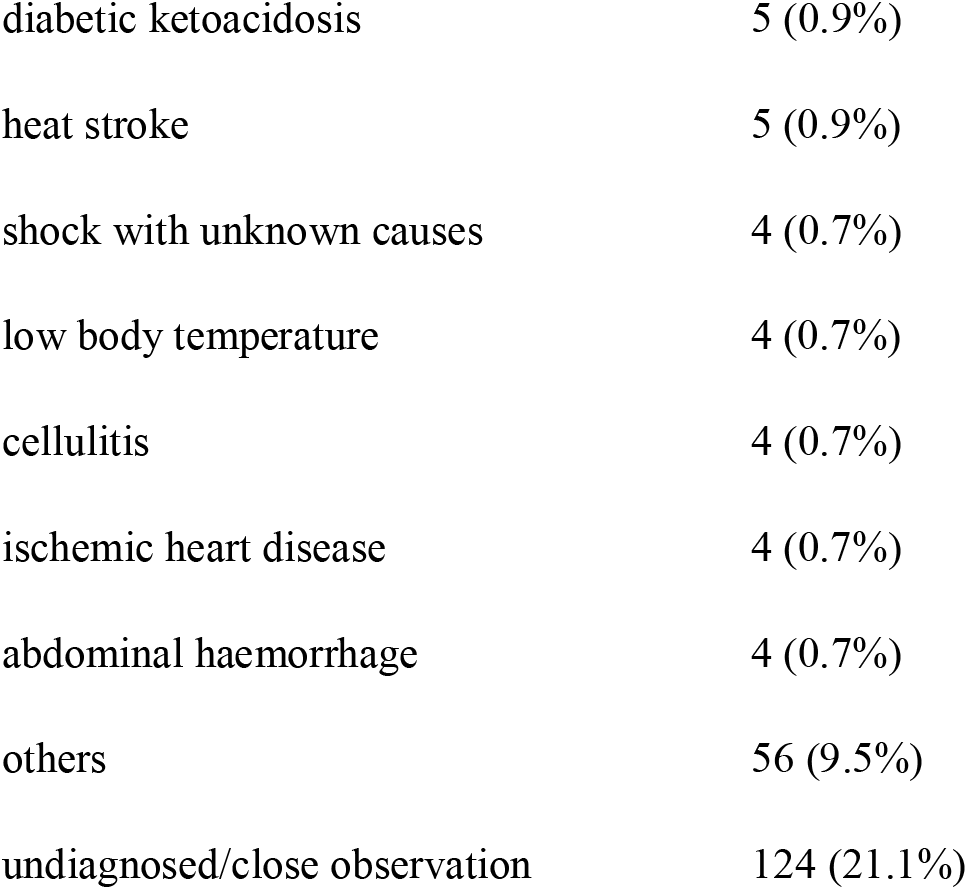
Non-stroke patients’ characteristics

The patient characteristics are shown in **Table 2**. We selected the most appropriate statistic test according to the distribution of each variable, as mentioned in the Methods section. Systolic pressure was significantly higher in the stroke group than in the non-stroke group (171.2 mmHg and 130.3 mmHg, respectively). The SI was significantly lower in the stroke group than in the non-stroke group (0.514 and 0.748, respectively). The other variables were not significantly different in these three groups.

**Table 2.**
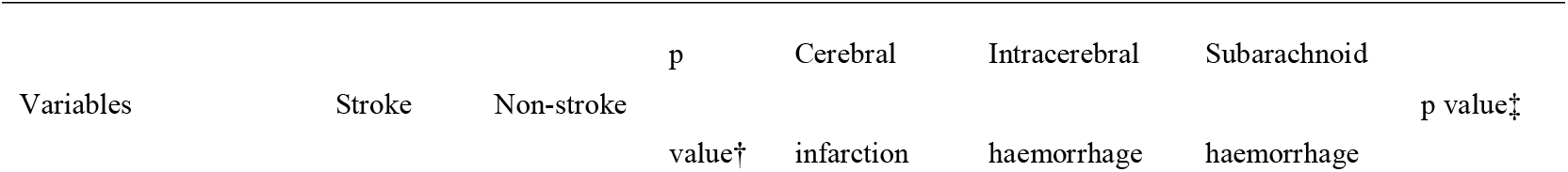
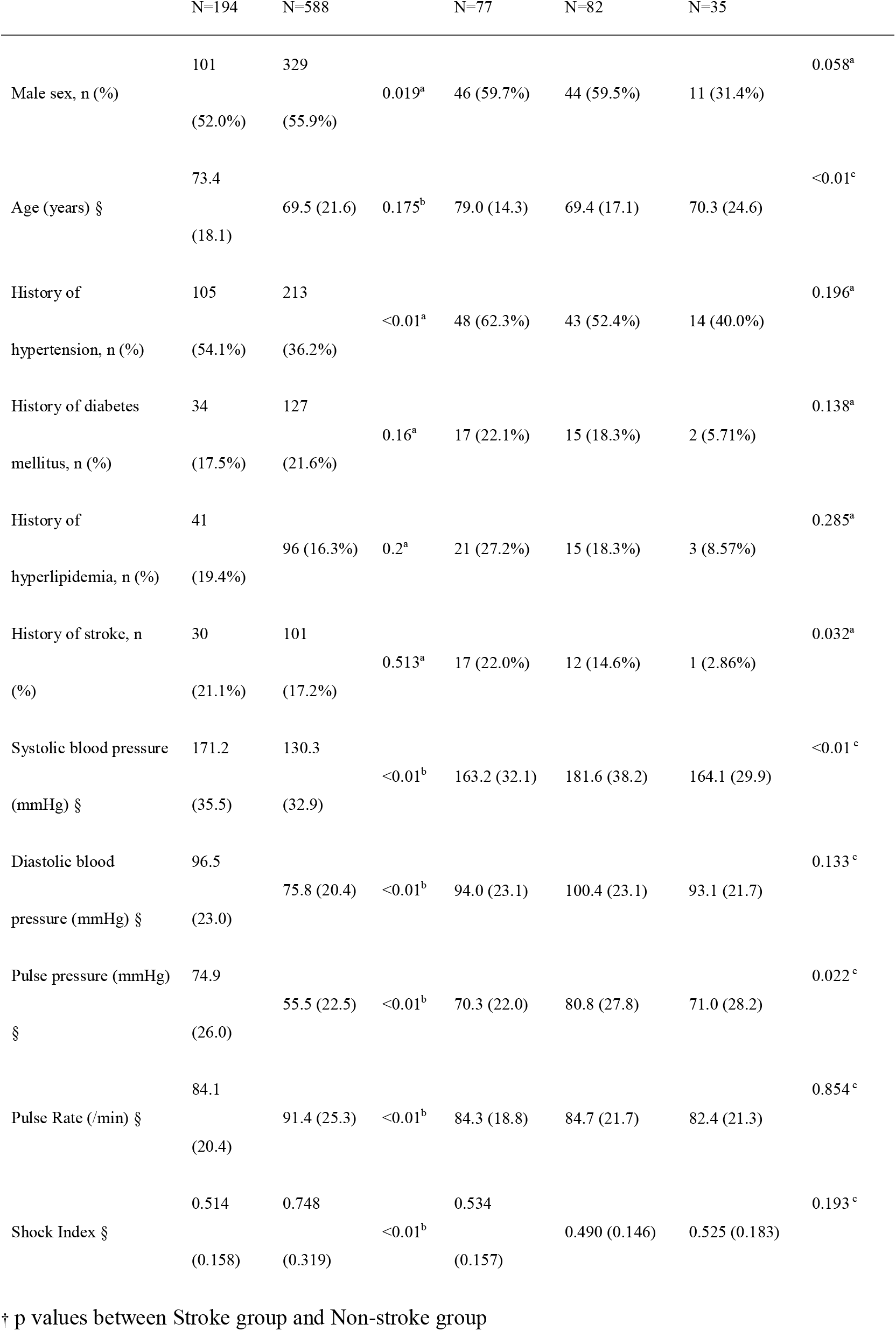
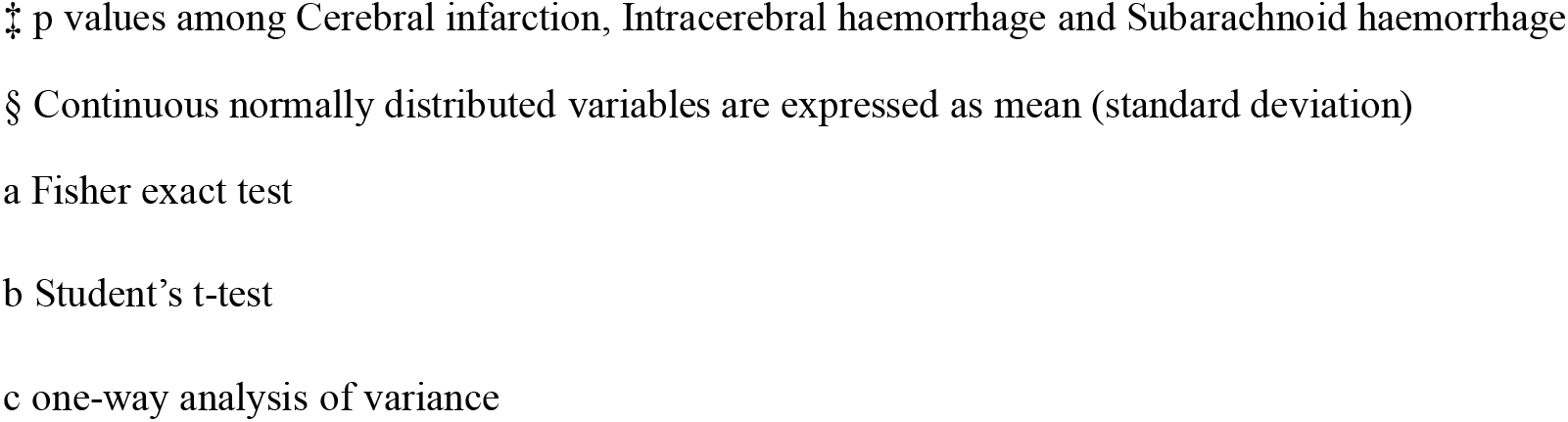
Patient characteristics

**Patient characteristics**
| Variables | Stroke | Non-stroke | p<br>value† | Cerebral<br>infarction | Intracerebral<br>haemorrhage | Subarachnoid<br>haemorrhage | p value‡ |
| --- | --- | --- | --- | --- | --- | --- | --- |

|  | N=194 | N=588 |  | N=77 | N=82 | N=35 |  |
| --- | --- | --- | --- | --- | --- | --- | --- |
| Male sex, n (%) | 101<br>(52.0%) | 329<br>(55.9%) | 0.019<sup>a</sup> | 46 (59.7%) | 44 (59.5%) | 11 (31.4%) | 0.058<sup>a</sup> |
| Age (years) § | 73.4<br>(18.1) | 69.5 (21.6) | 0.175<sup>b</sup> | 79.0 (14.3) | 69.4 (17.1) | 70.3 (24.6) | <0.01<sup>c</sup> |
| History of hypertension, n (%) | 105<br>(54.1%) | 213<br>(36.2%) | <0.01<sup>a</sup> | 48 (62.3%) | 43 (52.4%) | 14 (40.0%) | 0.196<sup>a</sup> |
| History of diabetes mellitus, n (%) | 34<br>(17.5%) | 127<br>(21.6%) | 0.16<sup>a</sup> | 17 (22.1%) | 15 (18.3%) | 2 (5.71%) | 0.138<sup>a</sup> |
| History of hyperlipidemia, n (%) | 41<br>(19.4%) | 96 (16.3%) | 0.2<sup>a</sup> | 21 (27.2%) | 15 (18.3%) | 3 (8.57%) | 0.285<sup>a</sup> |
| History of stroke, n (%) | 30<br>(21.1%) | 101<br>(17.2%) | 0.513<sup>a</sup> | 17 (22.0%) | 12 (14.6%) | 1 (2.86%) | 0.032<sup>a</sup> |
| Systolic blood pressure (mmHg) § | 171.2<br>(35.5) | 130.3<br>(32.9) | <0.01<sup>b</sup> | 163.2 (32.1) | 181.6 (38.2) | 164.1 (29.9) | <0.01<sup>c</sup> |
| Diastolic blood pressure (mmHg) § | 96.5<br>(23.0) | 75.8 (20.4) | <0.01<sup>b</sup> | 94.0 (23.1) | 100.4 (23.1) | 93.1 (21.7) | 0.133<sup>c</sup> |
| Pulse pressure (mmHg) § | 74.9<br>(26.0) | 55.5 (22.5) | <0.01<sup>b</sup> | 70.3 (22.0) | 80.8 (27.8) | 71.0 (28.2) | 0.022<sup>c</sup> |
| Pulse Rate (/min) § | 84.1<br>(20.4) | 91.4 (25.3) | <0.01<sup>b</sup> | 84.3 (18.8) | 84.7 (21.7) | 82.4 (21.3) | 0.854<sup>c</sup> |
| Shock Index § | 0.514<br>(0.158) | 0.748<br>(0.319) | <0.01<sup>b</sup> | 0.534<br>(0.157) | 0.490 (0.146) | 0.525 (0.183) | 0.193<sup>c</sup> |

† p values between Stroke group and Non-stroke group
‡ p values among Cerebral infarction, Intracerebral haemorrhage and Subarachnoid haemorrhage
§ Continuous normally distributed variables are expressed as mean (standard deviation)
a Fisher exact test
b Student's t-test
c one-way analysis of variance

### 3-2. Diagnostic performance of sBP and SI

The AUCs for sBP, SI, and pulse pressure were 0.801 (95% confidence interval: 0.761 – 0.840), 0.764 (0.729 – 0.799), and 0.717 (0.673 – 0.761), respectively (**Fig. 2A**). The AUC for sBP was higher compared to that for sBP and pulse pressure (p = 0.043 and <0.001, respectively, DeLong’s test).

**Fig 2.**
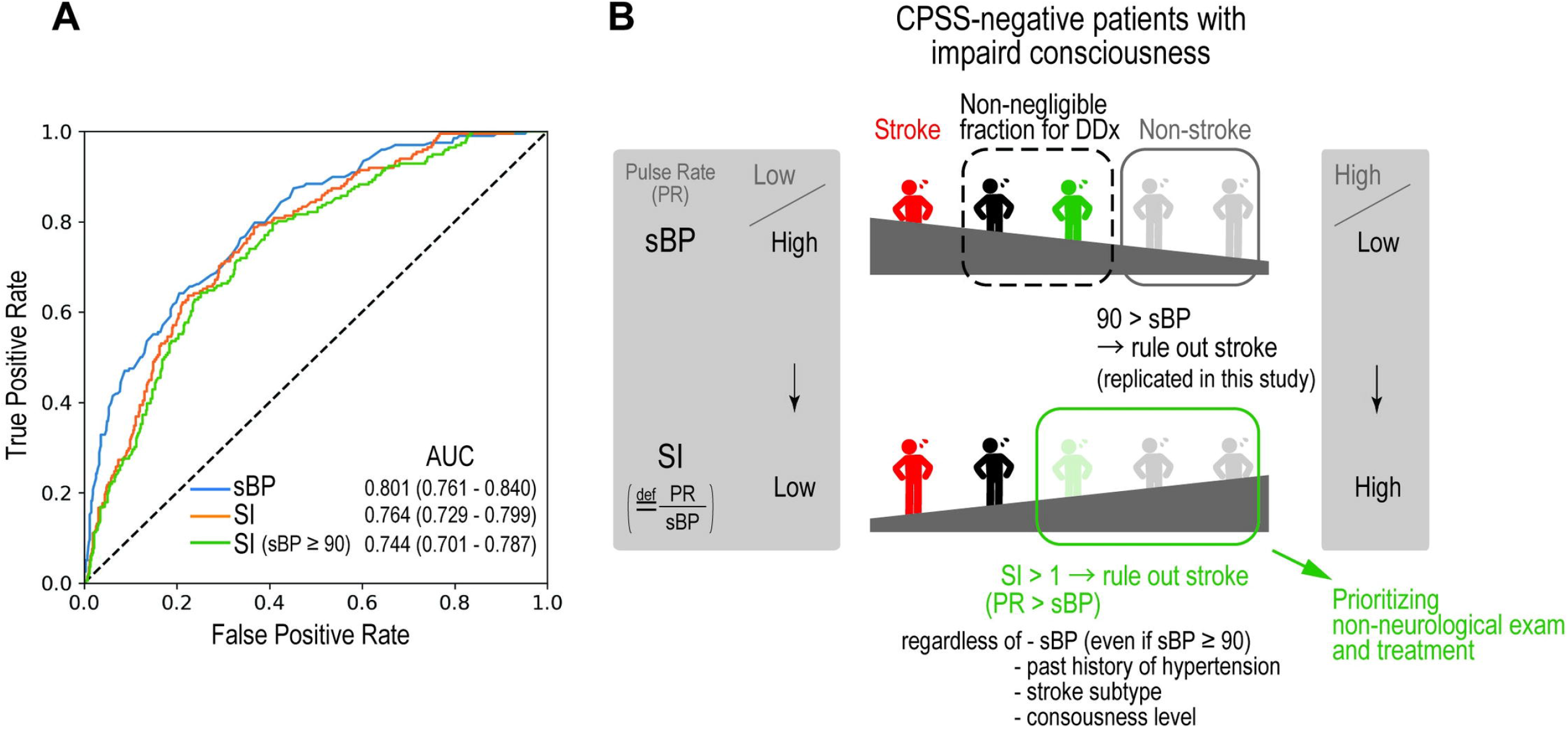
Diagnostic values of sBP and SI and relevance of the present study. (A) ROC curves and AUCs for sBP, SI, and SI in patients with sBP interval). AUC for sBP are higher than that of SI (p = 0.043, DeLong’s test). AUCs for SI and SI in patients with sBP ≥ 90 are not statistically different from each other (p = 0.44, DeLong’s test). (B) Relevance of the present study and proposal for clinical setting. Above: Stroke can be ruled out in patients with sBP lower than 90 mmHg (Gray persons); however, there is non-negligible fraction of patients who should be ruled out stroke (Black and Green persons). Bottom: When SI is higher than 1 (pulse rate exceeds sBP), stroke can be also ruled out based on the present study; therefore, the emergency physician can prioritize non-neurological examination and treatment. SI can be additionally applicable with sBP. See the main text for more details.

**Table 3** shows the sensitivity and specificity for each cut-off value of sBP and SI. Sensitivity for the cut-off values of sBP higher than 90 mmHg and SI lower than 1 reached 99.5% (95% confidence interval: 98.6 – 100%) and 99.5% (98.6 – 100%), respectively. With these cut-off values, specificity was 10.9% (9.61 – 12.2%) for an sBP of 90 mmHg and 15.1% (13.6 – 16.6%) for an SI of 1. The negative likelihood ratio for an sBP higher than 90 mmHg was 0.045 (0.038 – 0.052) and the one for an SI lower than 1 was 0.034 (0.029 – 0.039).

**Table 3.**
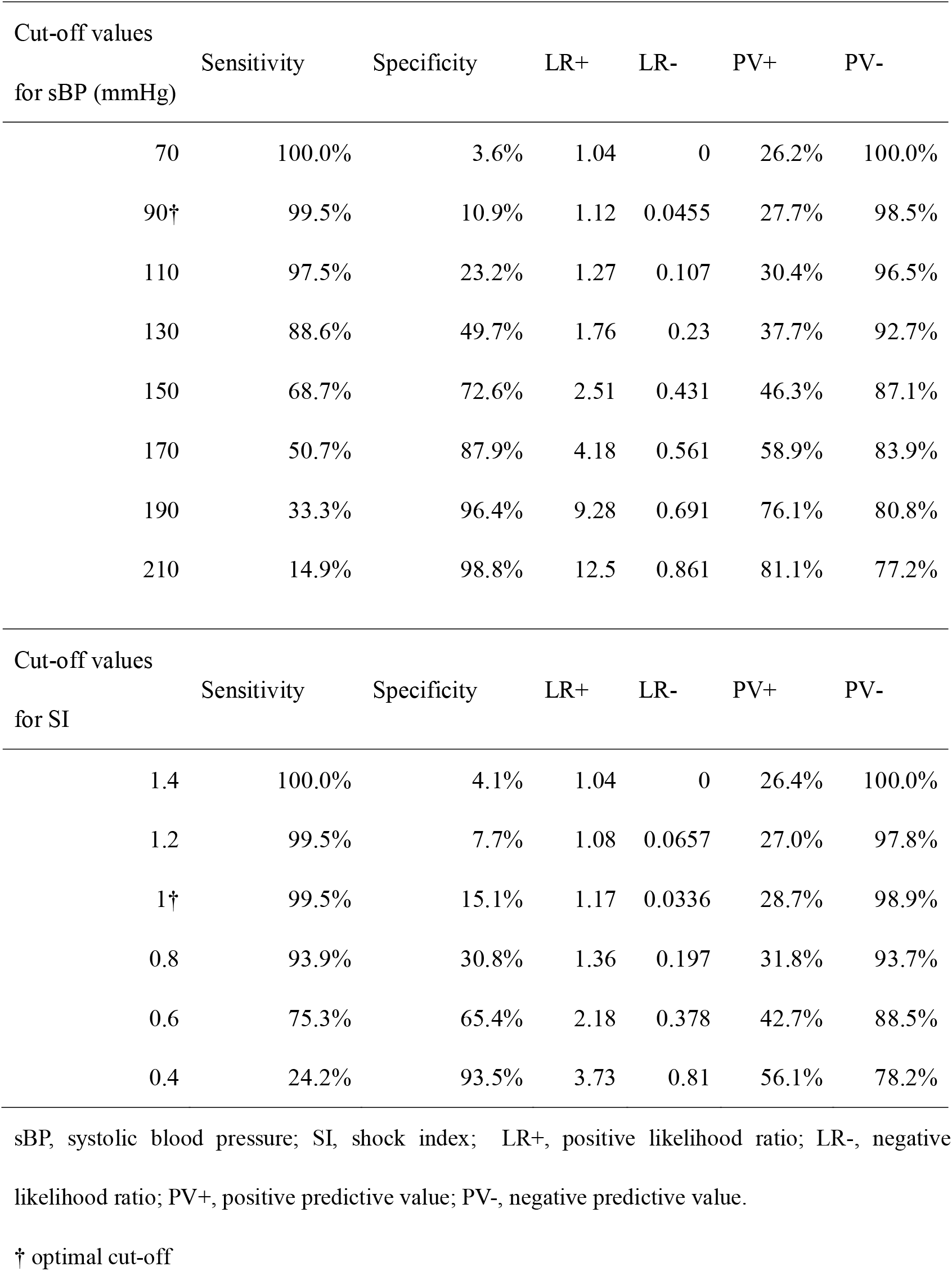
Cut-off values for systolic blood pressure and shock index for stroke

**Cut-off values for systolic blood pressure and shock index for stroke**
| Cut-off values<br>for sBP (mmHg) | Sensitivity | Specificity | LR+ | LR- | PV+ | PV- |
| --- | --- | --- | --- | --- | --- | --- |
| 70 | 100.0% | 3.6% | 1.04 | 0 | 26.2% | 100.0% |
| 90† | 99.5% | 10.9% | 1.12 | 0.0455 | 27.7% | 98.5% |
| 110 | 97.5% | 23.2% | 1.27 | 0.107 | 30.4% | 96.5% |
| 130 | 88.6% | 49.7% | 1.76 | 0.23 | 37.7% | 92.7% |
| 150 | 68.7% | 72.6% | 2.51 | 0.431 | 46.3% | 87.1% |
| 170 | 50.7% | 87.9% | 4.18 | 0.561 | 58.9% | 83.9% |
| 190 | 33.3% | 96.4% | 9.28 | 0.691 | 76.1% | 80.8% |
| 210 | 14.9% | 98.8% | 12.5 | 0.861 | 81.1% | 77.2% |

| Cut-off values<br>for SI | Sensitivity | Specificity | LR+ | LR- | PV+ | PV- |
| --- | --- | --- | --- | --- | --- | --- |
| 1.4 | 100.0% | 4.1% | 1.04 | 0 | 26.4% | 100.0% |
| 1.2 | 99.5% | 7.7% | 1.08 | 0.0657 | 27.0% | 97.8% |
| 1† | 99.5% | 15.1% | 1.17 | 0.0336 | 28.7% | 98.9% |
| 0.8 | 93.9% | 30.8% | 1.36 | 0.197 | 31.8% | 93.7% |
| 0.6 | 75.3% | 65.4% | 2.18 | 0.378 | 42.7% | 88.5% |
| 0.4 | 24.2% | 93.5% | 3.73 | 0.81 | 56.1% | 78.2% |

sBP, systolic blood pressure; SI, shock index; LR+, positive likelihood ratio; LR-, negative
likelihood ratio; PV+, positive predictive value; PV-, negative predictive value.
† optimal cut-off

### 3-3. Subgroup analysis: history of hypertension

We performed a subgroup analysis to determine whether a history of hypertension would affect the validity of these cut-off values. In patients with a past history of hypertension, the AUCs for sBP and SI were 0.752 and 0.729, respectively. For an sBP of 90 mmHg, the sensitivity and specificity were 99.0% and 7.21%, respectively, while for an SI of 1, they were at 99.0% and 7.69% respectively. In patients without a past history of hypertension, on the other hand, the AUCs for the sBP and SI were 0.822 and 0.780, respectively. The sensitivity and specificity for an sBP of 90 mmHg were 100% and 13.0%, respectively, while for an SI of 1 they measured 100% and 19.1%, respectively.

### 3-4. Subgroup analysis: consciousness level

We also performed another subgroup analysis to determine whether the validity of these cut-off values would be affected by the patient’s level of consciousness. Of 782 eligible patients, we had data available for 532 patients (68.0%) whose level of consciousness had been measured using the GCS. The other patients’ level of consciousness was recorded using the Japan Coma Scale, which is also commonly used in Japan. Of these 532 patients, 121 had stroke and 411 had non-stroke disease. **Table 4** shows the sensitivity and specificity of these cut-off values, both sBP higher than 90 and SI lower than 1, in the subgroups of patients with non-comatose level (GCS higher than 8), and comatose level (GCS less than or equal to 8). In the non-stroke group, the ratio of patients with SI equal to or higher than 1 is significantly higher in the subgroup of comatose level than in the subgroup of non-comatose level (p < 0.001, chi-squared test).

**Table 4.** Subgroup analysis of consciousness level (N = 532)

**Subgroup analysis of consciousness level (N = 532)**
|  | sBP > 90 |  | SI < 1 |  |
| --- | --- | --- | --- | --- |
|  | Sensitivity | Specificity | Sensitivity | Specificity |
| GCS > 8<br>(N = 365) | 98.5% | 13.1% | 100.0% | 13.1% |
| GCS ≤ 8<br>(N = 167) | 98.1% | 15.0% | 98.1% | 22.1% |

### 3-5. Sensitivity analysis

For sensitivity analysis, we evaluated the diagnostic performance of SI in different subgroups of sBP (**Table 5**). In the subgroup with sBP of 91–110, the specificity was higher, which indicated that SI would be better at ruling out stroke. In the subgroup of patients with sBP equals to or higher than 90, the AUC for SI was 0.744 (95% confidence interval: 0.701 – 0.787, **Fig. 2A**), which was not statistically different from AUC for SI in all patients (p = 0.44, DeLong’s test).

**Table 5.** Sensitivity and specificity of shock index lower than 1 in the subgroups of systolic blood pressure

**pressure**
| sBP | Number of patients | Sensitivity | Specificity |
| --- | --- | --- | --- |
| - 90 | 75 | 0.0% | 67.1% |
| 91 - 110 | 75 | 100.0% | 35.2% |
| 111 - 130 | 173 | 100.0% | 10.9% |
| 131 - 150 | 176 | 100.0% | 3.7% |
| 151 - 170 | 125 | 100.0% | 1.1% |
| 171 - | 170 | 100.0% | 0.0% |

## 4. Discussion

### 4-1. Relevance of the study

This study demonstrated that both the sBP and SI were useful in ruling out stroke in patients with impaired consciousness (**Table 3** and **Fig. 2A**). Previous studies showed the utility of sBP in diagnosing stroke [4–6]. According to one previous study, the positive likelihood ratio for an sBP higher than 170 mmHg was 6.09, and the negative likelihood ratio for an sBP lower than 90 mmHg was 0.04 [4]. Our study showed similar results of 4.18 and 0.045, respectively, verifying the diagnostic value of sBP in identifying patients with stroke.

Our study suggested that SI was another novel parameter that could be useful for ruling out stroke. Calculation of accurate SI is time-consuming and unsuitable for emergency settings; therefore, we suggest the cut-off value of 1. It means that a simple comparison of pulse rate and systolic blood pressure is enough to apply in the clinical setting. The probability of stroke would be sufficiently low in patients with an SI higher than 1, in other words, a pulse rate higher than their sBP (**Table 3**).

Moreover, SI can be used concurrently with sBP. As mentioned above, stroke can be ruled out in patients with sBP lower than 90 mmHg [4]; however, there is non-negligible fraction of patients who should be ruled out stroke. We showed that SI was still a useful indicator even in the patients with sBP equals to or higher than 90 (**Table 5** and **Fig. 2A**).

When SI was higher than 1 (pulse rate exceeded sBP), the emergency physician might be able to prioritize non-neurological examination and treatment, such as antibiotic administration and correction of electrolytes or blood sugar, over cranial imaging studies (**Fig. 2B**), since non-stroke causes of impaired consciousness include infection, electrolyte imbalance, and metabolic disorders (**Table 1**).

To control for possible confounding variables, we performed some subgroup analyses—subtype of stroke, level of consciousness, and past history of hypertension. We found that SI was not significantly different among the three sub-groups of patients with CI, ICH, or SAH, though the mean sBP was higher in the ICH group (**Table 3**). We showed that SI had good diagnostic performances regardless of level of consciousness (**Table 4**) and past history of hypertension, although the specificity could be low in the patients with a past history of hypertension. Taken together, we propose that stroke can be reasonably ruled out in more patients using both sBP and SI as indicators (**Fig. 2B**).

In Japan, CT scans and MRI are readily available in many tertiary care centres, including our institution. Once stroke is suspected, imaging is done to confirm the diagnosis [14]. In the present study, all eligible patients received the diagnosis of stroke based on CT and MRI results. Hence, the final diagnosis was definite in each case. However, we need to note that of all the patients with impaired consciousness who underwent a cranial CT scan, only 25.6% were diagnosed with stroke. Based on our results, we might be able to reduce unnecessary cranial CT scans in the future.

In cases of cardioembolic stroke, due to the possible background of atrial fibrillation, the pulse rate is expected to exceed the sBP. However, we experienced no cases of cardioembolic stroke with an SI higher than 1 in our study. We hypothesized that patients with cardioembolic stroke in the present study were appropriately screened out using the CPSS. Further studies are necessary to evaluate whether our results can be applied to patients with an irregular pulse.

### 4-2. Limitations and future directions

There were some limitations to this study. First, this was a single-centre review of patient records, with only a small number of cases of impaired consciousness. Second, we did not include patients who did not undergo a CT scan. However, as mentioned above, almost all patients experiencing a disturbance of consciousness underwent CT scan at our hospital. Third, our judgment as to the final diagnosis was based on the description of the physician in charge, which might have lacked objectivity and depended on his or her diagnostic skills. Fourth, in recent years, it has been pointed out that the amount of time elapsed since the vital sign was “last seen normal” is of importance to interpret vital signs. As we had no access to these data in this study, this is a potential confounding factor. Further large-scale, multi-centre, prospective studies are required to verify the utility of SI for diagnosing stroke. In addition, as the specificity of SI was not high enough, examinations with higher specificity, including CT, are still required to make the final diagnosis. Rapid and easily accessible measurement with higher specificity is desirable.

## 5. Conclusion

SI could serve as an indicator for stroke in CPSS-negative patients with impaired consciousness. Our study suggested that an SI higher than 1 ruled out stroke in a substantial percentage of adult patients with impaired consciousness. In cases in which the patient’s pulse rate is higher than their sBP, the emergency physician may be able to prioritize non-neurological examinations and treatments over cranial imaging studies (**Fig. 2B**).

## Data Availability

All relevant data are within the paper

## Acknowledgements

We thank Mr. James Robert Valera for his assistance in editing this manuscript, all of the staff and patients who contributed to this study.

## Abbreviations

AUC: area under the ROC curve
CPSS: Cincinnati Prehospital Stroke Scale
DDx: differential diagnosis
PR: pulse rate [/min]
ROC: receiver operating characteristic
SI: shock index
sBP: systolic blood pressure [mmHg]

## References

1. Kothari R, Barsan W, Brott T, Broderick J, Ashbrock S. Frequency and accuracy of prehospital diagnosis of acute stroke. Stroke. 1995;26: 937–941. doi:10.1161/01.str.26.6.937

2. Tamune H, Kuki T, Kashiyama T, Uchihara T. Does This Adult Patient With Jolt Accentuation of Headache Have Acute Meningitis? Headache. 2018;58: 1503–1510. doi:10.1111/head.13376

3. Jauch EC, Saver JL, Adams, HPJ, Bruno A, Connors, JJB, Demaerschalk BM, et al. Guidelines for the early management of patients with acute ischemic stroke: a guideline for healthcare professionals from the American Heart Association/American Stroke Association. Stroke. 2013;44: 870–947. doi:10.1161/STR.0b013e318284056a

4. Ikeda M, Matsunaga T, Irabu N, Yoshida S. Using vital signs to diagnose impaired consciousness: cross sectional observational study. BMJ. 2002;325: 800. doi:10.1136/bmj.325.7368.800

5. Geetadevi Y, Joshi R, Pai M, Kalantri SP. Simple clinical predictors of brain lesions in patients with impaired consciousness: a cross sectional study from a rural, tertiary hospital in central India. Clin Neurol Neurosurg. 2005;108: 25–31. doi:10.1016/j.clineuro.2004.12.018

6. Irisawa T, Iwami T, Kitamura T, Nishiyama C, Sakai T, Tanigawa-Sugihara K, et al. An association between systolic blood pressure and stroke among patients with impaired consciousness in out-of-hospital emergency settings. BMC Emerg Med. 2013;13: 24. doi:10.1186/1471-227X-13-24

7. Bott R. Guyton and Hall Textbook of Medical Physiology 13ed. Igarss 2014. 2014. doi:10.1007/s13398-014-0173-7.2

8. Rady MY, Nightingale P, Little RA, Edwards JD. Shock index: a re-evaluation in acute circulatory failure. Resuscitation. 1992;23: 227–234. doi:10.1016/0300-9572(92)90006-x

9. Mutschler M, Nienaber U, Munzberg M, Wolfl C, Schoechl H, Paffrath T, et al. The Shock Index revisited – a fast guide to transfusion requirement? A retrospective analysis on 21,853 patients derived from the TraumaRegister DGU. Crit Care. 2013;17: R172. doi:10.1186/cc12851

10. Sankaran P, Kamath A V, Tariq SM, Ruffell H, Smith AC, Prentice P, et al. Are shock index and adjusted shock index useful in predicting mortality and length of stay in community-acquired pneumonia? Eur J Intern Med. 2011;22: 282–285. doi:10.1016/j.ejim.2010.12.009

11. McCall SJ, Musgrave SD, Potter JF, Hale R, Clark AB, Mamas MA, et al. The shock index predicts acute mortality outcomes in stroke. Int J Cardiol. 2015;182: 523–527. doi:10.1016/j.ijcard.2014.12.175

12. Ono K, Wada K, Takahara T, Shirotani T. Indications for computed tomography in patients with mild head injury. Neurol Med Chir (Tokyo). 2007;47: 291–298. doi:10.2176/nmc.47.291

13. Kanda Y. Investigation of the freely available easy-to-use software “EZR” for medical statistics. Bone Marrow Transplant. 2013;48: 452–458. doi:10.1038/bmt.2012.244

14. Redd V, Levin S, Toerper M, Creel A, Peterson S. Effects of fully accessible magnetic resonance imaging in the emergency department. Acad Emerg Med. 2015;22: 741–749. doi:10.1111/acem.12686

